# Surveillance of SARS-CoV-2 lineage B.1.1.7 in Slovakia using a novel, multiplexed RT-qPCR assay

**DOI:** 10.1101/2021.02.09.21251168

**Authors:** Viera Kováčová, Kristína Boršová, Evan D Paul, Monika Radvánszka, Roman Hajdu, Viktória Čabanová, Monika Sláviková, Martina Ličková, Ľubomíra Lukáčiková, Andrej Belák, Lucia Roussier, Michaela Kostičová, Anna Líšková, Lucia Maďarová, Mária Štefkovičová, Lenka Reizigová, Elena Nováková, Peter Sabaka, Alena Koščálová, Broňa Brejová, Edita Staroňová, Matej Mišík, Tomáš Vinař, Jozef Nosek, Pavol Čekan, Boris Klempa

## Abstract

**Background:** The emergence of a novel SARS-CoV-2 variant of concern called B.1.1.7 lineage sparked global alarm due to evidence of increased transmissibility, mortality, and uncertainty about vaccine efficacy, thus accelerating efforts to detect and track the variant. Current approaches to detect lineage B.1.1.7 include sequencing and RT-qPCR tests containing a target assay that fails or results in reduced sensitivity towards the B.1.1.7 variant.

**Aim:** Since many countries lack robust genomic surveillance programs and failed assays detect multiple unrelated variants containing similar mutations as B.1.1.7, we sought to develop an RT-qPCR test that can accurately and rapidly differentiate the B.1.1.7 variant from other SARS-CoV-2 variants.

**Methods:** We used bioinformatics, allele-specific PCR, and judicious placement of LNA-modified nucleotides to develop a test that differentiates B.1.1.7 from other SARS-CoV-2 variants. We validated the test on 106 clinical samples with lineage status confirmed by sequencing and conducted a surveillance study of B.1.1.7 lineage prevalence in Slovakia.

**Results:** Our multiplexed RT-qPCR test showed 97% clinical sensitivity at detecting lineage B.1.1.7. The assay was used in a country-wide surveillance of B.1.1.7 lineage spread in Slovakia. Retesting nearly 7,000 SARS-CoV-2 positive samples obtained during three campaigns performed within a one month period, revealed pervasive spread of B.1.1.7 with an average prevalence of 82%.

**Conclusion:** Labs can easily implement this test to rapidly scale B.1.1.7 surveillance efforts and it is particularly useful in countries with high prevalence of variants possessing only the ΔH69/ΔV70 deletion because current strategies using target failure assays incorrectly identify these as putative B.1.1.7 variants.

## Introduction

In Dec 2020, Rambaut et al. [1] reported the genomic characterization of a distinct phylogenetic cluster named lineage B.1.1.7 (also referred to as 20I/501Y.V1 by Nextstrain (https://nextstrain.org/sars-cov-2/) or Variant of Concern (VOC) 202012/01), briskly spreading over the past four weeks in the United Kingdom. The new lineage has 23 mutations: 13 non-synonymous mutations, 4 deletions, and 6 synonymous mutations. The spike protein contains ten mutations at the amino-acid level (ΔH69/ΔV70 and ΔY144 deletions, N501Y, A570D, D614G, P681H, T716I, S982A, D1118H) that could potentially change binding affinity of the virus [2–4] and consequently virus-host interaction. Indeed, emerging evidence suggests lineage B.1.1.7 has enhanced transmissibility [3, 5–8], results in higher viral loads [9, 10], and causes increased mortality [11]. These data highlight the need for tools to facilitate enhanced surveillance of lineage B.1.1.7 as well as other variants that may harbour spike gene mutations that alter viral dynamics.

The dominant approach used to putatively detect lineage B.1.1.7 involves conducting multigene RT-qPCR tests that result in positive detection of SARS-CoV-2 in one or more gene sets together with a so-called S gene target failure (SGTF), which is used as a proxy for the B.1.1.7 variant. This approach permits widespread, rapid screening, but is limited by the fact that other variants, in addition to lineage B.1.1.7, produce SGTFs. Thus, the SGTF screening method depends on the presence of other variants in a region and how they vary over time. Whole genome sequencing is the gold-standard for detection of the B.1.1.7 variant. This provides direct confirmation of the variant and identification of emerging variants; however, it is expensive, time consuming, low throughput, and many countries lack a robust genomic surveillance program, making this approach unwieldy to adopt for tracking and mitigating the spread of the B.1.1.7 variant.

Here, we report the development of a novel, multiplexed RT-qPCR test for differentiating lineage B.1.1.7 from all other SARS-CoV-2 lineages. We validated the test using a selected set of 106 clinical samples collected during routine testing with the lineage status verified by sequencing. Unlike other tests that rely on indirect detection via SGTF, this test contains primers that target the ΔH69/ΔV70 and ΔY144 deletions in the spike gene that permit the direct detection of lineage B.1.1.7. This assay was used in three rounds of a country-wide screening of the prevalence of B.1.1.7 in Slovakia, which included 6,886 SARS-CoV-2 positive samples, revealing increasing prevalence of lineage B.1.1.7 over time in Slovakia. This RT-qPCR assay provides a useful tool for countries to rapidly identify hot spots of this new B.1.1.7 variant and implement test, trace, and isolate strategies to prevent this variant from becoming widespread. Countries currently experiencing extensive circulation of variants carrying only the ΔH69/ΔV70 deletion may find this test particularly useful as these would be falsely identified as the B.1.1.7 variant via SGTF tests.

## Materials and Methods

### Identification of RT-qPCR targets by bioinformatic analysis

To identify suitable targets for primer/probe design, we downloaded 1,136 sequences from the GISAID repository filtered during a collection time spanning 1 - 21 December 2020. We focused on the spike gene because lineage B.1.1.7 contains a number of spike gene mutations, including two deletions (ΔH69/ΔV70 and ΔY144) that are ideal for designing a specific assay. We cut the locus encoding the spike protein and used the MAFFT alignment tool (with the parameter - auto)[12] to align all the sequences against the WUHAN reference (NCBI ID: NC_045512.2). Twelve sequences (1.06 %) contained ambiguous signal in the loci of deletions and were not used in the downstream analysis. We separated sequences into two groups: 1) those with the ΔH69/ΔV70 and ΔY144 deletions and 2) those without the deletions (**Supplementary Table S1**). Using SeaView [13], we called 95% consensus sequences for the ΔH69/ΔV70 and ΔY144 group and the No deletions group that were subsequently used to design primer and probe sets specific to either B.1.1.7 or all other SARS-CoV-2 variants, respectively.

**Table 1.**
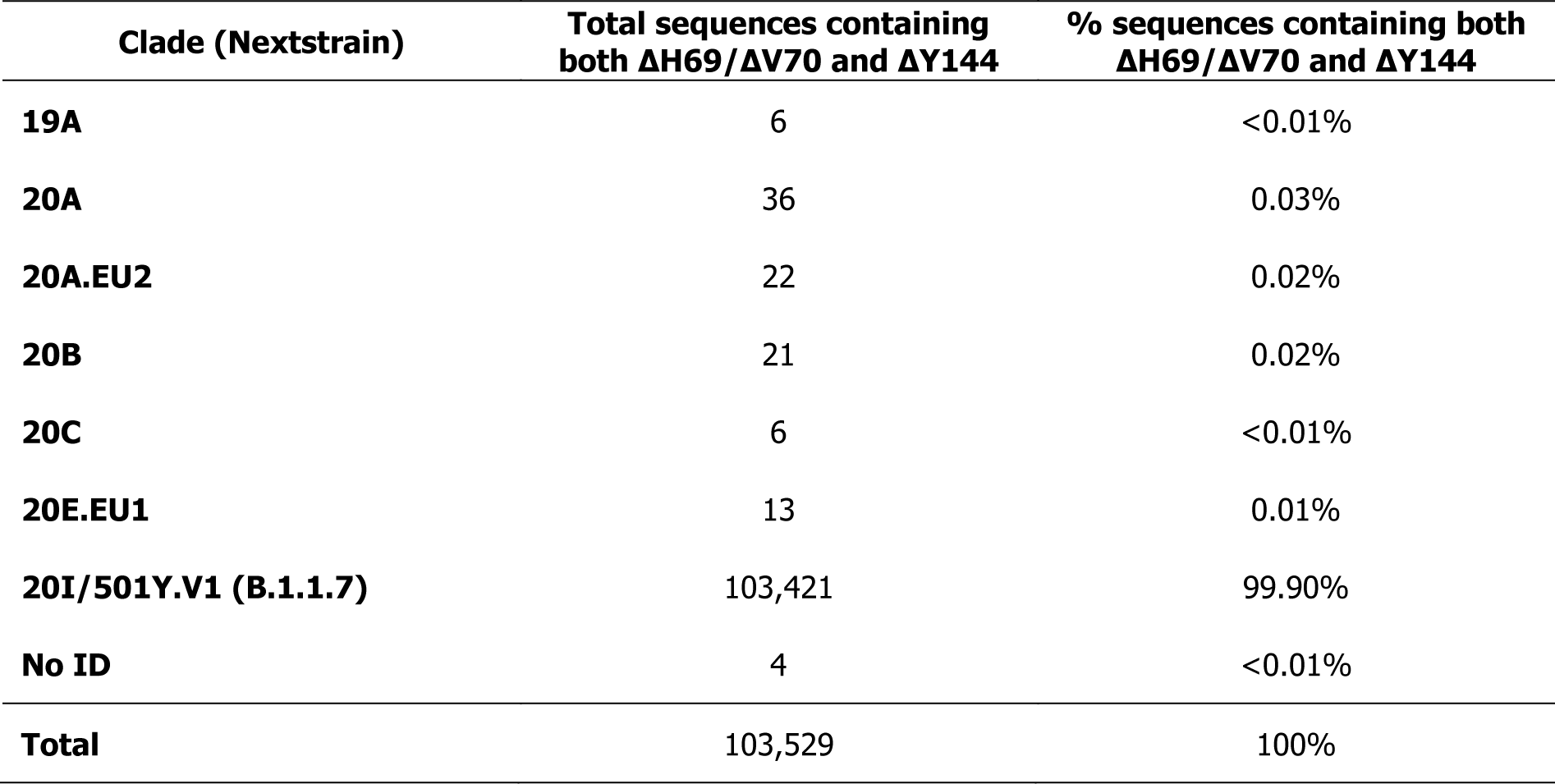
Summary of GISAID sequences containing both Spike ΔH69/ΔV70 and ΔY144 deletions

In a separate analysis to determine the prevalence of the ΔH69/ΔV70 and ΔY144 deletions in lineages other than B.1.1.7, we downloaded 633,137 spike protein sequences with the most recent data description file collected from the beginning of the pandemic through 2 March 2021. Using regular expressions (bash pattern matching command grep with the option -P for Perl-compatible regular expression), we searched for loci with both ΔH69/ΔV70 and ΔY144 deletions and for loci without these deletions. In the regular expression, we kept fixed a few amino acids downstream and upstream from the deletions to omit any miscalling of the searched pattern. All commands and scripts are available here: https://github.com/MultiplexDX/B117-RT-qPCR-design/blob/main/B1117.ipynb.

### Primer design and synthesis

We designed primers and probes using the 95% consensus sequences to target the S gene of the common SARS-CoV-2 (called SARS-CoV-2 S gene assay). To differentiate the B.1.1.7 variant from all other SARS-CoV-2 variants, we also designed primers and probes to target the S gene of SARS-CoV-2 variants containing either the ΔH69/ΔV70 deletion or the ΔY144 deletion, or both deletions (called B.1.1.7 assay). As an internal control, we synthesized a primer/probe set for human RNase P published by the US CDC [14]. We incorporated locked nucleic acid (LNA)-modified bases into some primers and probes following general guidelines in order to normalize melting temperatures, increase sensitivity, and enhance specificity [15–17]. Following primer/probe design, we conducted *in silico* analyses using the IDT OligoAnalyzer™ tool (https://www.idtdna.com/pages/tools/oligoanalyzer) to verify melting temperature (Tm), GC content, and potential to form homo-/hetero-dimers as well as the mFold server [18] (http://www.bioinfo.rpi.edu/applications/mfold/) to identify problematic secondary structures or necessary hairpin formation for TaqMan probes. Probes for both SARS-CoV-2 S gene and B.1.1.7 were labelled with a 5’-FAM (6-carboxyfluorescein) reporter dye and 3’-BHQ-1 (black hole quencher 1) dye. In multiplexed reactions, the probe for human RNase P was labelled with a 5’-Cy5 (cyanine 5) reporter dye and 3’-BHQ-3 dye. Primers and probes were synthesized at MultiplexDX, Inc. (Bratislava, Slovakia; https://www.multiplexdx.com/). The sequences of primers and probes used in this study are listed in **Supplementary Table S2**. This test is also available as an *in vitro* diagnostic (IVD) CE marked kit called rTest COVID-19 B.1.1.7 qPCR kit (https://www.multiplexdx.com/products/rtest-covid-19-b-1-1-7-qpcr-kit, MultiplexDX, Inc.).

**Table 2.**
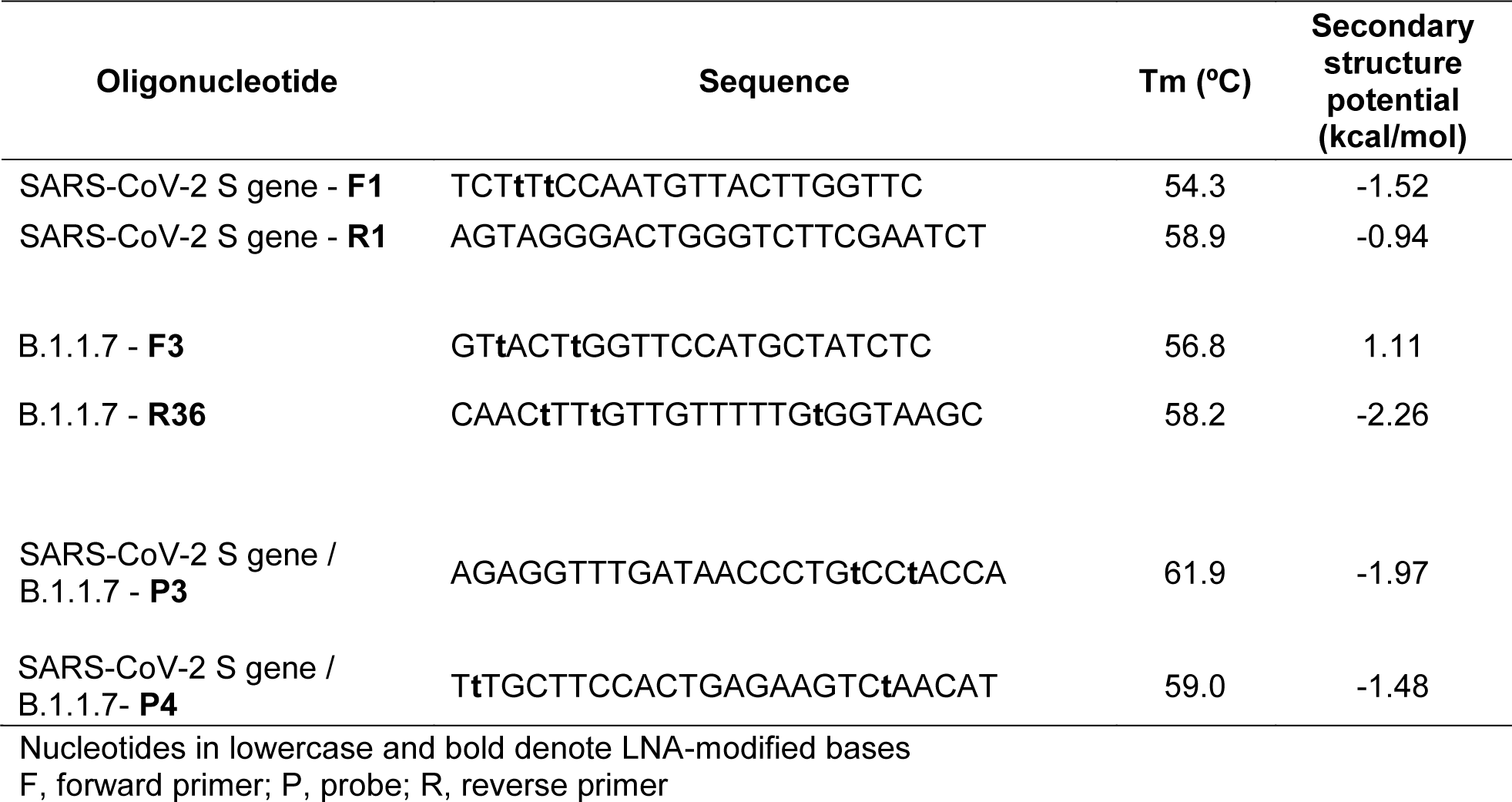
Oligonucleotide primers and probes for common SARS-CoV-2 S gene and B.1.1.7 primer/probe sets

### Positive controls

For primer/probe set optimization, we used the following positive controls: 1) RNA extracted from a patient confirmed positive for a common variant of SARS-CoV-2 that does not contain any deletions in the spike gene (named wild type template), 2) RNA extracted from a patient confirmed positive for a common variant of SARS-CoV-2 that contains the six base pair deletion (bp: 21765-21770) resulting in the deletion of two amino acids at the 69/70 position of the spike protein (named ΔH69/ΔV70 template), and 3) RNA extracted from a patient confirmed positive by whole genome sequencing for the SARS-CoV-2 lineage B.1.1.7 (named B.1.1.7 template). The SARS-CoV-2 control samples were confirmed by whole genome sequencing of tiled ∼ 2-kbp long amplicons on a MinION device (Oxford Nanopore Technologies, Oxford, UK) essentially as described by Resende *et al*. (2020) [19].

### RT-qPCR

We optimized RT-qPCR reactions and conducted clinical validations using both an AriaMx (Agilent, CA, USA) and QuantSudio 5 (ThermoFisher Scientific, MA, USA) real-time PCR system. For all the detected genes, we used the SOLIScript® 1-step CoV Kit (Cat. No. 08-65-00250, SOLIS BioDyne, Tartu, Estonia) according to the manufactureŕs recommendations comprised of 4 µl of 5X One-step Probe CoV Mix (ROX), 0.5 µl of 40X One-step SOLIScript® CoV Mix, 2 µl of primers/probe mix, 8.5 µl of PCR water, and 5 µl of sample in a 20 µl total volume. One-step RT-qPCR assays were conducted with the following cycling conditions: 55 °C for 10 min for reverse transcription, 95 °C for 10 min for initial denaturation, and 45 cycles of 95 °C for 15 s and 60 °C for 30 s. Concentrations for primers and probes were as follows: SARS-CoV-2 S gene (forward and reverse primer = 500 nM, probes = 200 nM for each probe (single and dual); B.1.1.7: forward primer = 600 nM, reverse primer = 800 nM, probes = 200 nM for each probe (single and dual); RNase P (forward and reverse primer = 250 nM, probe = 80 nM).

### Analytical sensitivity (limit of detection)

To assess the analytical sensitivity of both our common SARS-CoV-2 S gene (S gene) and B.1.1.7 primer/probe sets, we used RNA isolated from a patient sample infected with the B.1.1.7 variant of SARS-CoV-2 as confirmed by sequencing. This RNA was diluted to 200 copies/μl and then serial dilutions were prepared by diluting the stock with a synthetic matrix “SARS-CoV-2 Negative“ (Cat. No. COV000, Exact Diagnostics, TX, USA) containing genomic DNA at a concentration of 75,000 copies/ml, resulting in samples with concentrations of 8 copies/μl (= 40 copies/reaction), 2 copies/μl (= 10 copies/reaction), 0.8 copies/μl (= 4 copies/reaction), 0.4 copies/μl (= 2 copies/reaction) and 0.2 copies/μl (= 1 copy/reaction) that were used in the analytical sensitivity test. The assay was performed in 8 replicates for each prepared dilution.

### Clinical performance evaluation

We evaluated the clinical utility of our SARS-CoV-2 S gene and B.1.1.7 primer/probe sets using a selected set of 106 clinical samples, which were confirmed by an RT-qPCR reference method used for routine testing by regional public health authorities of the Slovak Republic. Further sequencing revealed 67 of these samples belonging to the B.1.1.7 lineage, 24 samples belonging to the B.1.258 lineage (contains only the ΔH69/ΔV70 deletion), and 15 samples belonging to other SARS-CoV-2 lineages. The SARS-CoV-2 sequences were determined by sequencing of tiled ∼ 2-kbp long amplicons on a MinION device (Oxford Nanopore Technologies, Oxford, UK) as described by Resende et al. (2020) [19].

We conducted a detailed assessment of potential biases and applicability judgements of the clinical validation using the QUADAS-2 tool [20] (http://www.bristol.ac.uk/population-health-sciences/projects/quadas/quadas-2/; **Supplementary Table S3**). Inclusion criteria for the selected set of samples consisted of a positive result from a reference standard RT-qPCR test as well as available sequencing information to confirm lineage. No SARS-CoV-2 negative samples were included in the clinical evaluation because in practice this test was to be used only as a tool to identify lineage status in samples previously identified as SARS-CoV-2 positive. The clinical validation was conducted by the Biomedical Research Center, Institute of Virology, Slovak Academy of Sciences (BMC-SAS). BMC-SAS received clinical samples previously identified as SARS-CoV-2 positive by local laboratories, then extracted RNA and performed the index test (rTest COVID-19 qPCR ALLPLEX KIT; MultiplexDX) and the B.1.1.7 test in parallel. All samples were processed and tested in a timely manner to minimize the effects of RNA degradation. For the clinical evaluation, researchers used a prespecified criterion to interpret test results (**Supplementary Table S4**) and were blind to the sequencing outcome.

**Table 3.**
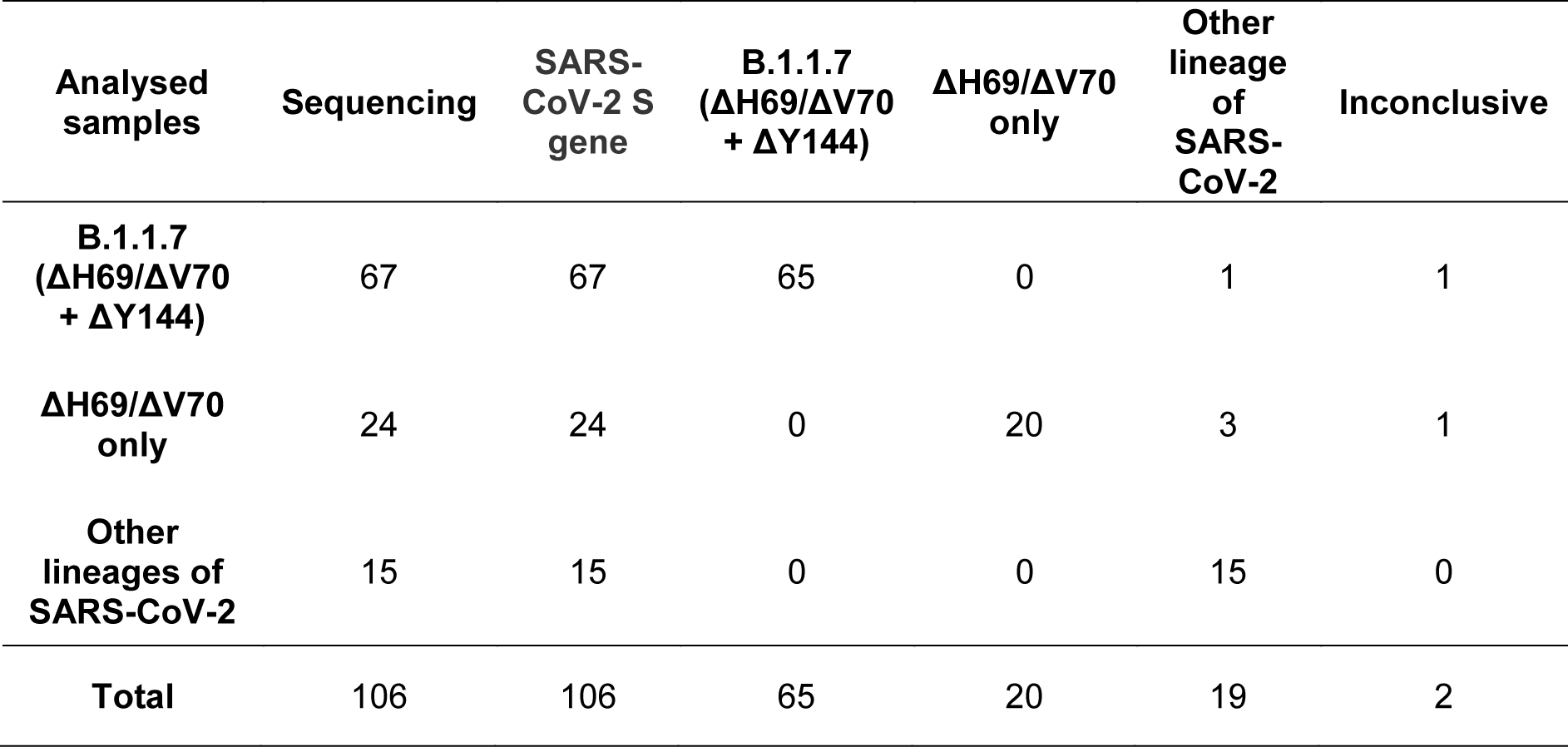
Clinical performance of SARS-CoV-2 S gene and B.1.1.7 primer/probes sets.

### Surveillance of lineage B.1.1.7 prevalance throughout the Slovak Republic

This test (rTest COVID-19 B.1.1.7 qPCR kit) was used to assess the prevalence of B.1.1.7 throughout the Slovak Republic over a period of one month. Multiple laboratories of the Public Health Authority of the Slovak Republic retested 6,886 samples that were identified as SARS-CoV-2 positive by a standard RT-qPCR test. Retesting occurred over a one month period on February 2^nd^, 2021 (1,962 samples), February 17^th^, 2021 (2,382 samples), and March 3^rd^, 2021 (2,542 samples). To ensure sufficient sample size for smaller geographic regions, we grouped districts based on the jurisdictions of the regional public health offices (there are 36 public health offices each covering 1-8 districts) and distributed tests accordingly. Laboratory personnel determined lineage status using the predefined criteria outlined in **Supplementary Table S4**. For comparison wth our observed B.1.1.7 prevalances, we also used the prevalences obtained on the first screening round (February 2^nd^, 2021) to estimate historical and future prevalences using a range of two weeks and applying spread factors (3.3 and 5.4) that were derived from estimated reproduction numbers from the UK [5, 8]. Data from the surveillance screening can be found on GitHub: https://github.com/Institut-Zdravotnych-Analyz/covid19-data/tree/main/PCR_Tests.

## Results

### Identification of RT-qPCR targets by bioinformatic analysis

Our analysis of 1,136 spike gene sequences (spanning 1 - 21 December 2020) revealed 228 sequences (20%) that contained both the ΔH69/ΔV70 and ΔY144 deletions (for country of origin, see **Supplementary Table S1**). The shorter deletion (ΔY144) always co-occurred with the longer deletion (ΔH69/ΔV70), whereas the (ΔH69/ΔV70) deletion occurs independently in 17 sequences (1.5%). Pearson’s correlation coefficient of the deletions is 0.953.

We analysed over 600,000 SARS-CoV-2 genomes to determine the prevalence of both ΔH69/ΔV70 and ΔY144 deletions in lineage B.1.1.7 and lineages other than B.1.1.7 and found a total of 103,529 sequences that possess both deletions. Based on the metadata file, we identified SARS-CoV-2 lineages across all called sequences with both deletions. Only 108 sequences (0.10%) out of 103,529 sequences are not labelled as B.1.1.7. In other words, 99.90% of sequences containing both deletions belong to lineage B.1.1.7, highlighting the notion that these two deletions are highly specific for the B.1.1.7 variant and make ideal targets for primer/probe design (**Table 1**).

### Development of a multiplexed RT-qPCR assay to distinguish lineage B.1.1.7 from all other SARS-CoV-2 variants

To develop a multiplexed RT-qPCR test to distinguish lineage B.1.1.7, we designed two assays targeting either the wild-type SARS-CoV-2 spike gene or the ΔH69/ΔV70 and ΔY144 deletions that are highly specific to lineage B.1.1.7 (for primer/probe locations and sequences see **Figure 1** and **Table 2**, respectively).

**Figure 1.**
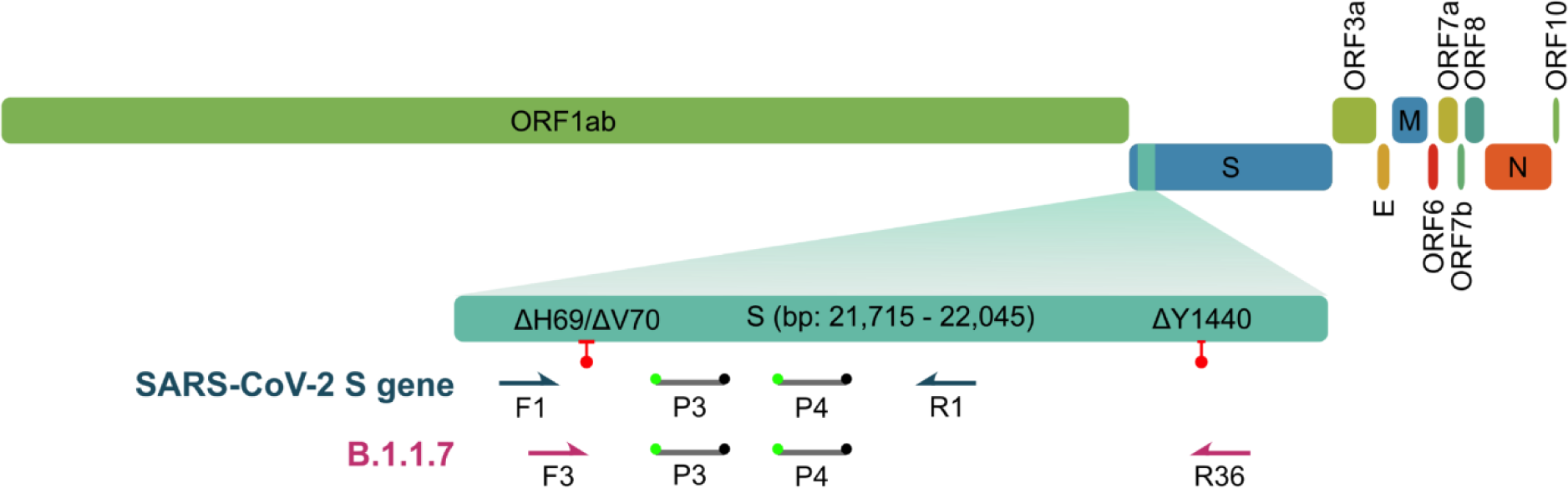
Development and optimization of a general SARS-CoV-2 S gene primer/probe set for all SARS-CoV-2 variants. Schematic illustrates genomic organization of SARS-CoV-2 with an emphasis on the location of both SARS-CoV-2 S gene and B.1.1.7 primer/probe sets relative to the ΔH69/ΔV70 and ΔY144 deletions (red lines with ball points) observed in the spike gene of B.1.1.7 variants.

We began by designing a general S gene primer/probe assay (SARS-CoV-2 S gene) that could be used for screening purposes and would detect the most common strains of SARS-CoV-2 as well as variants containing the ΔH69/ΔV70, including lineage B.1.1.7. We assessed the performance of a series of primers flanking the ΔH69/ΔV70 deletion (**Supplementary Figure S1A, B**; for primer/probes sequences, see **Supplementary Table S2.**). After selecting the optimal primer/probe combination, we introduced an additional hydrolysis probe, identically labelled with the same reporter and quencher dyes, that would hybridize in tandem (i.e., on the same strand) with the first hydrolysis probe. Consistent with other reports [21, 22], this dual probe approach enhanced sensitivity and specifity (**Supplementary Figure S1C, D**).

For our assay targeting lineage B.1.1.7, we leveraged the highly specific co-occurrence of the ΔH69/ΔV70 and ΔY144 deletions in lineage B.1.1.7 (99.90%, **Table 1**). By designing a series of forward primers to target the ΔH69/ΔV70 deletion, we differentiated wild type template from ΔH69/ΔV70 template (**Supplementary Figure S2A**). Since other SARS-CoV-2 variants share the ΔH69/ΔV70 deletion (e.g., B.1.1.298, B.1.160, B.1.177, B.1.258, B.1.375, B.1.525), we designed a series of reverse primers to target the second, three base pair deletion (bp: 21991-21993; ΔY144) and utilized allele-specific PCR approaches [23–25] and judicious placement of LNA-modified nucleotides to enhance the specificity of the assay (**Supplementary Figure S2B** and **C**). This approach enabled us to differentiate B.1.1.7 variants that contain both the ΔH69/ΔV70 and ΔY144 deletions from SARS-CoV-2 variants that contain only the ΔH69/ΔV70 deletion, provided that a second reaction is ran in parallel using the SARS-CoV-2 S gene set that can be used as a benchmark to assess the relative sensitivity. If the B.1.1.7 primer set amplifies the sample within five Ct cycles of the SARS-CoV-2 S gene primer set, then the sample is B.1.1.7 positive. Alternatively, if the B.1.1.7 primer set amplifies the sample in 8 or more Ct cycles relative to the SARS-CoV-2 S gene primer set, than the sample likely belongs to a variant that contains the ΔH69/ΔV70 deletion, but not the ΔY144 deletion, and hence is B.1.1.7 negative.

We compared three different versions of B.1.1.7 primer/probe sets using a selected set of 46 samples, some of which were sequenced to confirm lineage status. Given our interpretation criterion (**Supplementary Table S4**), we determined that the V3 primer/probe set performed the best since it correctly identified all B.1.1.7 and ΔH69/ΔV70 deletion samples, with the exception of one ΔH69/ΔV70 deletion sample that was interpreted as inconclusive (**Supplementary Figure S3**).

### Analytical sensitivity and clinical evaluation of lineage B.1.1.7 S gene primer/probe set

With our final primer/probe sets for SARS-CoV-2 S gene and B.1.1.7 (**Table 2**), we multiplexed each assay with the US CDC human RNase P primer/probe set (for sequences, see [14]) as an internal control to assess RNA extraction and assay performance. We then determined the analytical sensitivity using serial dilutions of RNA extracted from a B.1.1.7 positive sample. Both assays displayed high sensitivity (**Figure 2A**) with our SARS-CoV-2 S gene and B.1.1.7 assays reliably detecting down to only 2 copies/reaction (0.4 copies/μl) and 10 copies/reaction (2 copies/μl), respectively, placing them among the most sensitive SARS-CoV-2 RT-qPCR assays available.

**Figure 2.**
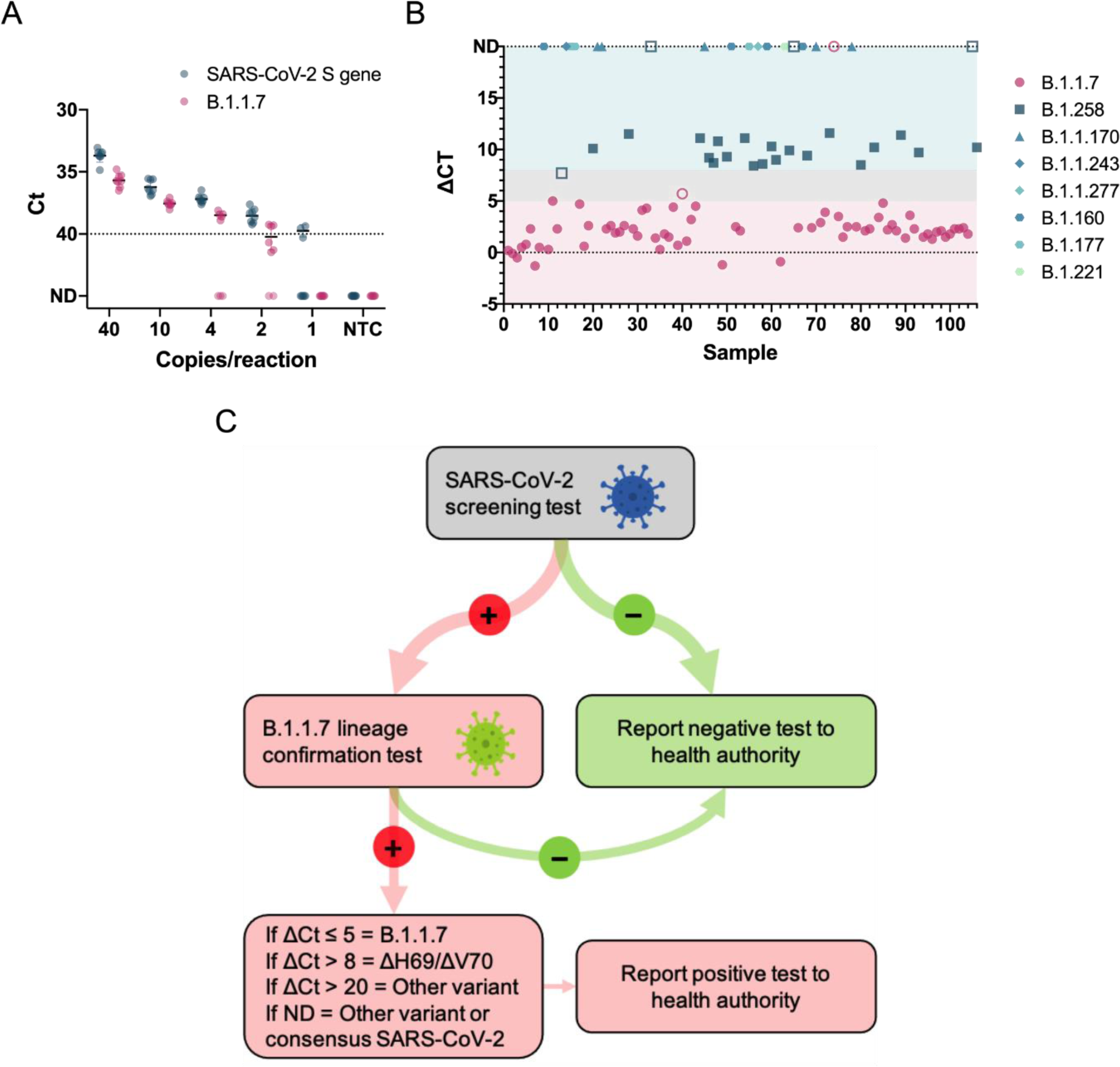
Analytical sensitivity and clinical validation of SARS-CoV-2 S gene and B.1.1.7 assays. **A)** The limit of detection was determined for both SARS-CoV-2 S gene and B.1.1.7 assays by serial dilutions of isolated viral B.1.1.7 RNA. Data depict the mean and SD of eight replicates per each dilution. The dotted line at Ct 40 serves as a threshold after which amplification is considered invalid. **B)** Overview of ΔCt values (= B.1.1.7 assay Ct − SARS-CoV-2 S gene assay Ct) for each sample in the clinical validation. Symbols represent the various SARS-CoV-2 lineages that were identified by sequencing. Closed symbols represent samples correctly identified by either the SARS-CoV-2 S gene or B.1.1.7 assays, whereas open symbols denote samples that did not meet the criterion established for variant identification. The shaded background shows ΔCt ranges that correspond with the criterion to report a sample as B.1.1.7 positive (pink), ΔH69/ΔV70 deletion positive (teal), and inconclusive (gray). ND, not detected. **C)** Decision tree demonstrating the proper workflow, interpretation criterion, and actions to implement the SARS-CoV-2 S gene and B.1.1.7 assays a testing regime to identify B.1.1.7 positive samples.

We evaluated the clinical performance of our SARS-CoV-2 S gene and B.1.1.7 assays on 106 clinical samples that underwent sequencing to identify lineage status using interpretation criterion outlined in **Supplementary Table S4**. Our SARS-CoV-2 S gene assay detected all 106 clinical samples regardless of lineage (**Table 3**) confirming its utility as a general screening assay for the most common SARS-CoV-2 variants. Out of 67 clinical samples classified as lineage B.1.1.7 by sequencing, our B.1.1.7 assay positively identified 65 samples, while one sample (Sample 75) was not detected by the B.1.1.7 assay and another sample was deemed inconclusive. The ΔCt of this sample was slightly greater than five cycles (e.g., Sample 40, ΔCt = 5.7) relative to the Ct value for the SARS-CoV-2 S gene assay, which exceeded our cut-off for a positive identification (**Figure 2B**).

Notably, our assay was also capable of identifying samples carrying the ΔH69/ΔV70 deletion such as those belonging to the B.1.258 lineage, provided that the sample contains sufficient viral load as other ΔH69/ΔV70 variants yield ΔCt values greater than 8 Ct cycles. For the 24 samples that carry only the ΔH69/ΔV70 deletion and belong to lineage B.1.258, our B.1.1.7 assay correctly identified 20 out of 24 samples. Three samples were not detected by the B.1.1.7 assay possibly due to relatively high Ct values in the SARS-CoV-2 S gene assay for two samples (Sample 33, Ct = 30.1 and Sample 65, Ct = 28.9), making confirmation of the ΔH69/ΔV70 status impossible with our cut-off criterion. One sample had a ΔCt outside the criterion for ΔH69/ΔV70 deletion confirmation and was deemed inconclusive. Overall, the clinical evaluation confirmed the diagnostic utility of both our SARS-CoV-2 S gene and B.1.1.7 assays, which showed 100% (106/106) and 97% (65/67) diagnostic sensitivity, respectively. The assay also showed diagnostic utility for identifying variants containing only the ΔH69/ΔV70 deletion by detecting 83.3% (20/24) of lineage B.1.258 samples. When considering B.1.258 samples containing high viral loads (Ct ≤ 28), which is necessary to identify variants with only the ΔH69/ΔV70 deletion, the diagnostic sensitivity reached 91.7% (22/24). For an overview of the clinical evaluation data, lineage of each sample, and GISAID information, see **Supplementary Table S5**. We have provided a decision tree (**Figure 2C**) that users may follow to implement this test to directly detect the presence of the B.1.1.7 variant.

### Surveillance of lineage B.1.1.7 reveals increasing prevalence throughout the Slovak Republic

To determine the prevalence of lineage B.1.1.7 and its spread throughout the Slovak Republic, regional public health authorities used this test to screen nearly 7,000 samples previously identified as SARS-CoV-2 positive during three screening rounds over a one month period. The first surveillance screening for lineage B.1.1.7 on February 2^nd^, 2021 revealed a region-specific prevalence of B.1.1.7 ranging from 52% (Žilina) to 85% (Trnava) with an overall B.1.1.7 prevalance of 75% (**Figure 3A-C**; **Supplementary Table S6**). During the second round of screening held on February 17^th^, 2021, the majority of regions (5 out of 8) showed increased prevalence, although due to reduced prevalence in Banská Bystrica, Košice, and Nitra, the overall prevalence remained at 74%. Closer scrutiny of the data suggests the reduced prevalence in some regions was caused by samples originating from several “non-B.1.1.7 clusters” (e.g., in social care homes), where all positive samples were likely derived from a common infection source. Exclusion of these clusters results in a slight increase in prevalence of 78%. A final screening on March 3^rd^. 2021 showed increased B.1.1.7 prevalence in all regions and an overall prevalence of 85%. Taking into consideration estimates of growth rates (i.e., reproduction number) for lineage B.1.1.7 in the UK [5, 8], we used the prevalences obtained on the first screening round (February 2^nd^, 2021) to estimate historical and future prevalences using a range of two weeks and applying spread factors (3.3 and 5.4) estimated in other countries (**Figure 3D**). Although the expected data did not fit with our observed data, a number of factors likely influence regional spread, including the presence and virulence of competing SARS-CoV-2 lineages and selection biases from cluster outbreaks.

**Figure 3.**
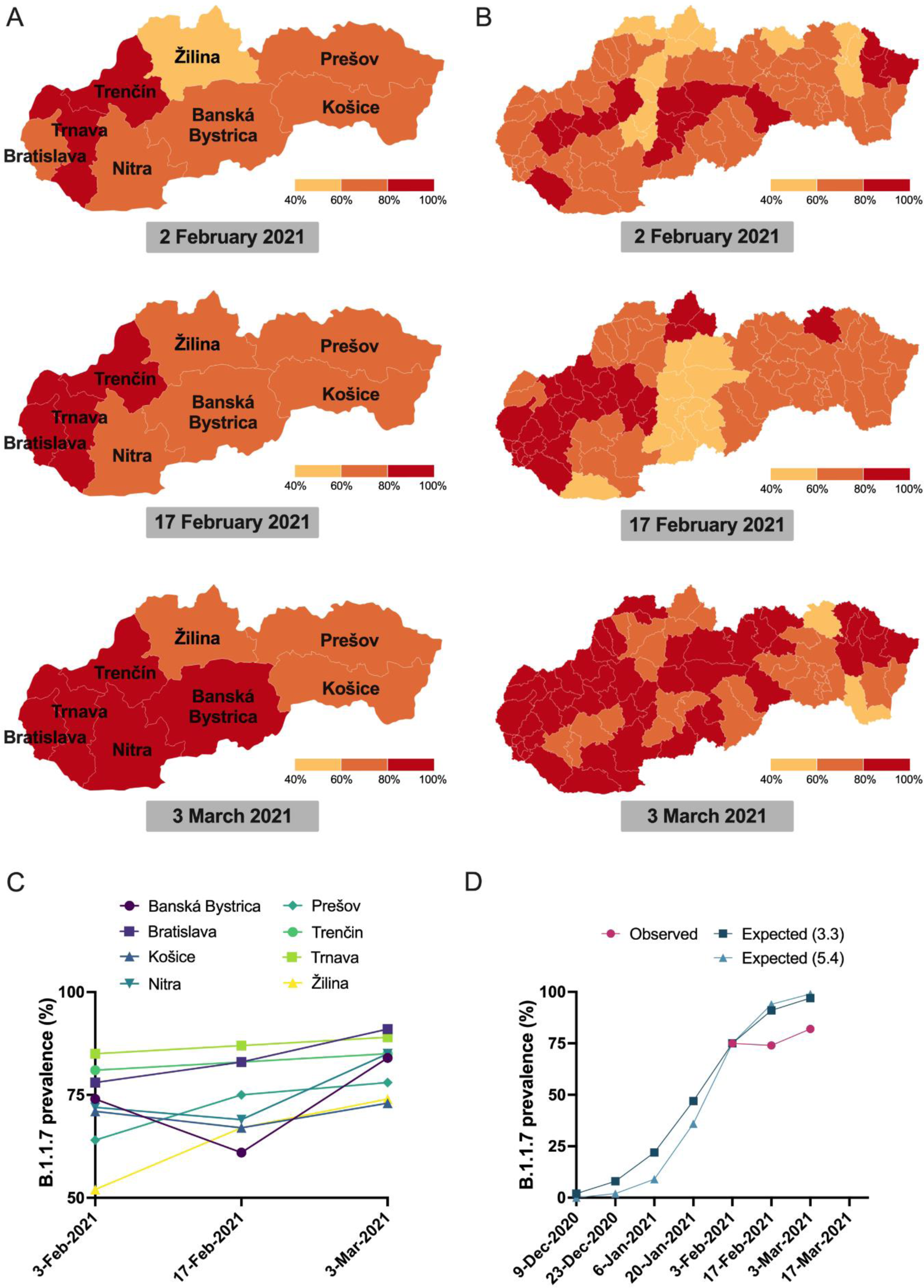
Tracking the prevalence of lineage B.1.1.7 in Slovakia. **A)** Maps of the prevalence of lineage B.1.1.7 in the eight regions of Slovakia during the three screening rounds held on February 2^nd^, 2021, February 17^th^, 2021, and March 3^rd^, 2021. Regions in red have prevalence > 80%, regions in yellow have prevalence <60%. **B)** Maps of the prevalence of lineage B.1.1.7 in the 79 districts of Slovakia during the three screening rounds held on February 2^nd^, 2021, February 17^th^, 2021, and March 3^rd^, 2021. Regions in red have prevalence > 80%, regions in yellow have prevalence <60%. **C)** Trends in B.1.1.7 prevalence during the three screening rounds in each of the eight regions of Slovakia. **D)** Observed trends in B.1.1.7 prevalence during the three screening rounds and expected prevalence given various two-week spread factors (shown in parentheses) assuming the prevalence of February 2^nd^, 2021.

## Discussion

The recent emergence of a novel SARS-CoV-2 variant called lineage B.1.1.7 has sparked global consternation as it has now been confirmed in over 70 countries and threatens to exacerbate an already dire pandemic. To mitigate the spread of the B.1.1.7 variant, it is imperative that countries have diagnostic tools that can quickly and accurately detect and track the prevalence of the variant in order to implement the appropriate epidemiological measures. Here we report a novel RT-qPCR test to differentiate the B.1.1.7 variant from other SARS-CoV-2 lineages. The test consists of running two S gene target assays, one specific for B.1.1.7 and the other for all SARS-CoV-2 strains, and performing a simple comparison of relative Ct values that allows the user to differentiate the B.1.1.7 variant from other variants that have the ΔH69/ΔV70 deletion. We validated this test on clinical samples that were sequenced to determine the exact SARS-CoV-2 lineage and the results demonstrated a high level of sensitivity in distinguishing the B.1.1.7. variant. Moreover, we retested nearly 7,000 samples previously confirmed as SARS-CoV-2 positive by a reference method in three screening rounds over a one month period, revealing widespread and increasing prevalence of lineage B.1.1.7 throughout the Slovak Republic. This RT-qPCR test allows a positive identification of the B.1.1.7 variant, providing countries with a powerful tool to detect and track lineage B.1.1.7, especially countries that have considerable prevalence of variants carrying only the ΔH69/ΔV70 deletion [26–29], which are mistakenly identified as B.1.1.7 variants by currently used SGTF assays.

Although there are hundreds of approved RT-qPCR tests for the detection of SARS-CoV-2, few of them are capable of directly differentiating the B.1.1.7 variant from common variants of SARS-CoV-2. Paradoxically, failed RT-qPCR tests have been instrumental in identifying putative B.1.1.7 positive samples and tracking its prevalence [26, 28, 30–33]. These RT-qPCR tests contain multigene assays, with at least one assay targeting the spike gene, and during routine testing a “drop-out” in the spike gene assay may occur (often termed as S gene target failure, or SGTF), while other gene targets yield positive signals. This SGTF can indicate the presence of the B.1.1.7 variant and flag samples for confirmation by sequencing. It is important to note, however, that SGTPs are produced by other variants that contain the ΔH69/ΔV70 deletion, including the B.1.1, B1.258, B.1.525, and B.1.1.298 (the mink cluster V) lineages, as well [7]. This highlights the importance of follow up sequencing of SGTF samples to determine lineage status.

Indeed, an analysis of SGTFs and corresponding sequencing data by Public Health England revealed that SGTF assays were poor proxies for the presence of B.1.1.7 in early October with only 3% of SGTFs assays positively identifying a B.1.1.7 variant. The SGTF assays only became useful proxies when the variant spread and became more dominant in late November when the assays then detected over 90% of the variant [34]. Others have reached a similar conclusion [5, 27, 28, 31, 32, 35], suggesting that the success of SGTF assays depends on the location, time, and frequency of other variants that contain the ΔH69/ΔV70 deletion. This is particularly problematic, since the SGTF assays are the least accurate at the time when the B.1.1.7 variant is at low prevalence, precisely the time when an accurate test is needed most in order to establish effective mitigation strategies. Our test outlined here makes significant strides in this effort by accurately differentiating the B.1.1.7 variant with a test that does not rely on a SGTF, thus providing a rapid, accurate test that eliminates the need to conduct expensive and laborious sequencing to confirm lineage status.

Besides the SGTF tests, a number of commercially available SARS-COV-2 variant tests are emerging and have been used as surveillance strategies to monitor the prevalence of variants such as B.1.1.7 [35–40]. These tests are largely based on classical SNP genotyping methods using either probe-based genotyping or melting curve analyses and typically focus on mutations that are shared between many variants (e.g., N501Y, E484K, and ΔH69/ΔV70 deletion), making it difficult to distinguish between variants unless running multiple SNP assays and then making complex comparisons of melt curves. Similar to SGTF tests, these approaches, while providing a rapid snapshot of the presence of SARS-CoV-2 variants, often require follow up genomic sequencing to identify the particular variant.

Several groups have described publicly available RT-qPCR protocols for detection of lineage B.1.1.7 that can be divided into SNP genotyping assays using either SYBR- [41] or probe-based [42] melting curve analyses, multiplexed probe-based RT-qPCR [43–46], and a combination of target drop-out tests [47]. While these open source RT-qPCR protocols offer cost-effective, rapid strategies to directly detect multiple SARS-CoV-2 variants, some are limited by only assessing a small number of mutations that preclude identification of specific variants and most have yet to be tested in real world surveillance scenarios.

To differentiate B.1.1.7, we took an alternative approach by targeting both the ΔH69/ΔV70 and ΔY144 deletions using allele-specific PCR methods combined with judicious placement of LNA oligonucleotides. Together, these modifications provided us with a primer/probe set that retained specificity for B.1.1.7 variants and reduced specificity to other variants containing only the ΔH69/ΔV70 deletion. To highlight the specificity of this assay, our analysis of all GISAID sequences containing both the ΔH69/ΔV70 and ΔY144 deletions revealed that a staggering 99.90% of all these sequences belong to lineage B.1.1.7, ensuring that users can have high confidence that a positive B.1.1.7 assay result is a true positive. Our test, instead of relying on target failures to identify putative variants, provides a positive signal in the presence of B.1.1.7 and ΔH69/ΔV70 deletion variants that can easily be differentiated by comparing their relative Ct values to a common SARS-CoV-2 S gene primer/probe set that serves as a benchmark.

We successfully monitored the dynamics of lineage B.1.1.7 prevalence by retesting nearly 7,000 SARS-CoV-2 positive samples in three successive testing campaigns in Slovakia. This mass surveillance effort provided invaluable information about the spread and prevalence of lineage B.1.1.7 without having to conduct expensive and time-consuming genomic sequencing. Although our observed data did not fit models of previously established replication numbers for lineage B.1.1.7 [5, 8], we attribute this discrepancy to region specific presence of other competing SARS-CoV-2 variants. Indeed, a recently described B.1.258 variant that was extensively circulating throughout the Czech Republic and Slovakia [29] contains mutations in the spike protein (N439K and ΔH69/ΔV70 deletion) that result in higher viral loads and increased transmissibility [7, 29, 48]. It is plausible that the reproduction number of B.1.1.7 is altered in a time and region-dependent manner that is associated with circulation of B.1.258.

The B.1.1.7 RT-qPCR assay described here was also used to screen for B.1.1.7 prevalence in 122 SARS-CoV-2 positive samples in the city of Trenčín, Slovakia in December 2020. While we observed 81-85% prevalence of B.1.1.7 in the region of Trenčín in February 2021, Brejová and colleagues [29] reported only 4% prevalence of B.1.1.7 and 41% prevalence of variants containing only the ΔH69/ΔV70 deletion back in December 2020. This highlights two key points: 1) when considering the B.1.1.7 prevalence in Trenčín in December 2020, the extrapolated historical data using estimated reproduction numbers matches our observed prevalence seen in Trenčín region on February 2^nd^, 2021 (**Figure 5C**). 2) SGTF tests utilized early on in a scenario like Trenčín would result in many false positive B.1.1.7 samples. Overall, these data provide real-world evidence of how this B.1.1.7 RT-qPCR assay can be used in mass surveillance screening to capture critical epidemiological information about the dynamics of B.1.1.7. It is also important to note that the outcome of the B.1.1.7 RT-qPCR assay-based screening and the observed trend is greatly in line with the sequence data from Slovakia available in the GISAID repository (https://www.gisaid.org/). From the clinical samples collected in Slovakia in February 2021, 83.8% (n=210) were identified as B.1.1.7 lineage while in the samples from March 2021, 97.1% (n=1336) belonged to the B.1.1.7 lineage.

We have provided interested users with the primer and probe sequences to implement this B.1.1.7 assay in their own laboratories with the hope this can rapidly scale the ability of countries to identify the B.1.1.7 variant and implement epidemiological measures to mitigate its spread. This test can provide labs with a powerful tool to directly confirm the presence of the B.1.1.7 variant in a sample previously determined SARS-CoV-2 positive by an approved screening test, thus avoiding the use of target gene failure assays that can be plagued with low specificity and obviating the need to conduct burdensome and costly genomic sequencing. This is particularly important for countries that are experiencing extensive circulation of variants harbouring only the ΔH69/ΔV70 deletion as current RT-qPCR assays that rely on SGTFs erroneously classify these samples as presumptive B.1.1.7 variants.

## Data Availability

All sequence data referred to in the manuscript are available at gisaid.org.

## Acknowledgements and funding statement

We gratefully acknowledge the authors from the submitting and originating laboratories that shared genetic sequencing data with the GISAID initiative. We acknowledge the contribution of the oligonucleotide synthesis and production team at MultiplexDX for synthesizing the primers and probes in this study, the laboratory staff of the Slovak regional public health authorities and diagnostic laboratories for screening samples during surveillance of B.1.1.7, and the team at the Institute for Healthcare Analyses for processing the surveillance data. Surveillance of lineage B.1.1.7 using MuliplexDX tests was funded by the Ministry of Health of Slovakia. This project was supported by the European Union’s Horizon 2020 research and innovation program [EVA-GLOBAL project, grant agreement number 871029] (BK) and grants from the Slovak Research and Development Agency: PP-COVID-20-0017 (BK) and PP-COVID-20-0116 (PC, BK).

## Author contributions

BK, PC, VK, KB, MM, and EDP conceptualized and planned the study. VK conducted bioinformatic analyses for primer/probe design and inclusivity. PC designed and synthesized primers/probes. MR, RH, KB, VC optimized primer/probe sets on positive controls and clinical samples. AB, LRo, MK, AL, LM, MS, LRe, EN, PS, AK collected and provided clinical samples and associated metadata. KB, VC, BB, JN, TV, performed genomic sequencing and bioinformatics to determine sample lineage status. BK, KB, VC, MS, ML, LL performed clinical validation and analysed clinical data. MM and ES collected and evaluated samples from the B.1.1.7 lineage surveillance and analysed the data. EDP, BK, PC, MM, ES, VK, MR, RH, KB analysed and interpreted the data and wrote the manuscript. All authors provided critical comments and feedback on the manuscript.

## Supplementary material

**Supplementary Figure S1.**
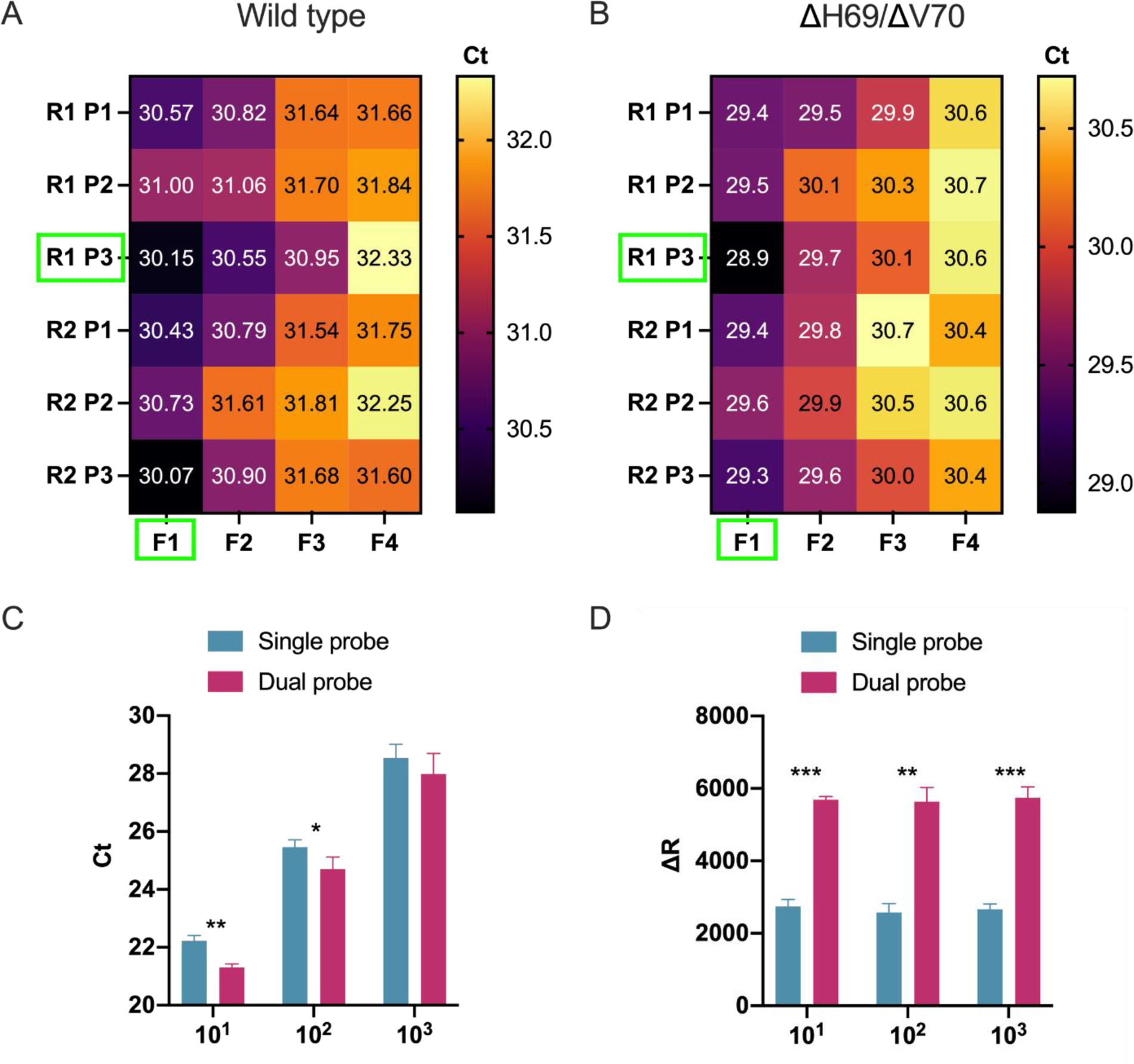
Development and optimization of a general SARS-CoV-2 S gene primer/probe set for all SARS-CoV-2 variants. A, B) Heatmaps illustrate oligonucleotide primer and probe combinations designed to target conserved sequences within the spike gene, including all SARS-CoV-2 variants that were contained in our bioinformatics analysis. Combinations of forward (F1-F4) and reverse primers (R1-R2) and hydrolysis probes (P1-P2) were tested using two separate SARS-CoV-2 variants, a common SARS-CoV-2 variant (Wild type, panel A) and a variant containing the ΔH69/ΔV70 deletion (B). Green rectangle boxes indicate best performing primer/probe combinations. C, D) Bar graphs compare RT-qPCR performance of a single probe versus an additional identically labelled dual probe using three 10-fold (10^1^, 10^2^, 10^3^) dilutions of SARS-CoV-2 template ran in triplicates. Evaluation of the performance was done by comparing raw Ct values (C) and fluorescence intensity values (D). Statistical analysis was performed using paired t-test (***p ≤ 0.001, **p ≤ 0.01, *p ≤ 0.05).

**Supplementary Figure S2.**
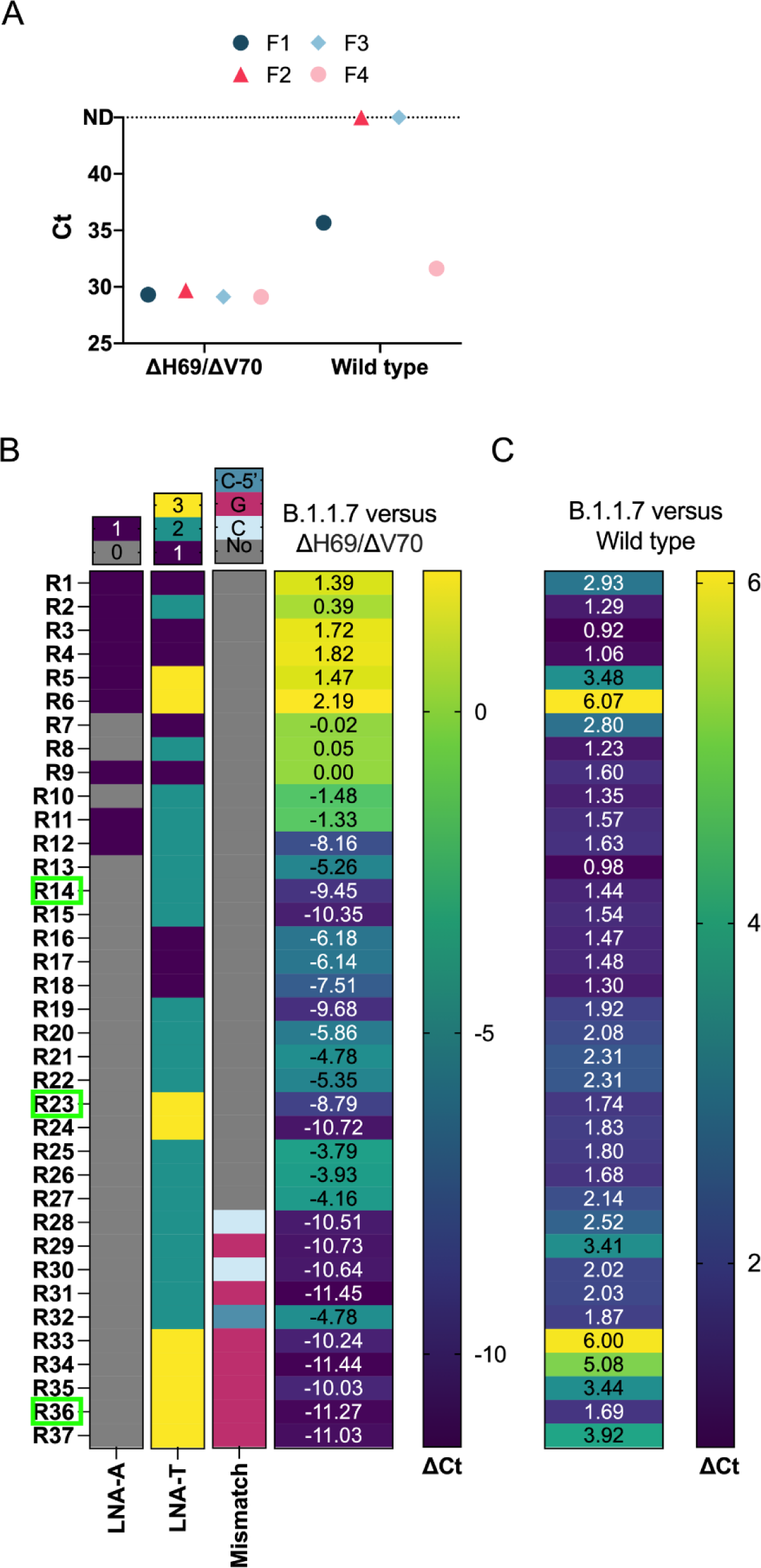
Development and optimization of a spike gene primer/probe set specific for the B.1.1.7 SARS-CoV-2 variant. A) Assessment of forward primers (F1-4) targeting the ΔH69/ΔV70 deletion in B.1.1.7 using the best reverse primer and probe (from **Supplementary Figure S1**/ **Supplementary Table S2**). Symbols compare Ct values of the ΔH69/ΔV70 variant and wild type templates. Dotted line indicates samples that were not detected (ND) within 45 cycles. B) Overview of reverse primer designs targeting the ΔY144 deletion of the B.1.1.7 variant and their effects on specificity by comparing the relative ΔCt when amplifying either B.1.1.7 or ΔH69/ΔV70 variants as template. Darker colours in the heatmap represent a greater ΔCt and consequently better specificity. Green rectangle boxes indicate reverse primers selected for further optimization. LNA-A depicts primers containing an LNA modified adenine base located at either the 3’- or 5’-end of the reverse primer. LNA-T displays the number (1-3) of LNA-modified thymine bases for each reverse primer. Mismatch base represents design modifications to introduce either a guanine (G) or cytosine (C) mismatch base in either the penultimate base (G or C) or the 3^rd^ from last base (C-5’) relative to the 3’-end of the reverse primer. C) Heatmap shows ΔCt value comparison of B.1.1.7 primer/probe set to SARS-CoV-2 S gene primer/probe set using the B.1.1.7 variant as template. Darker colours indicate smaller ΔCt and consequently better specificity.

**Supplementary Figure S3.**
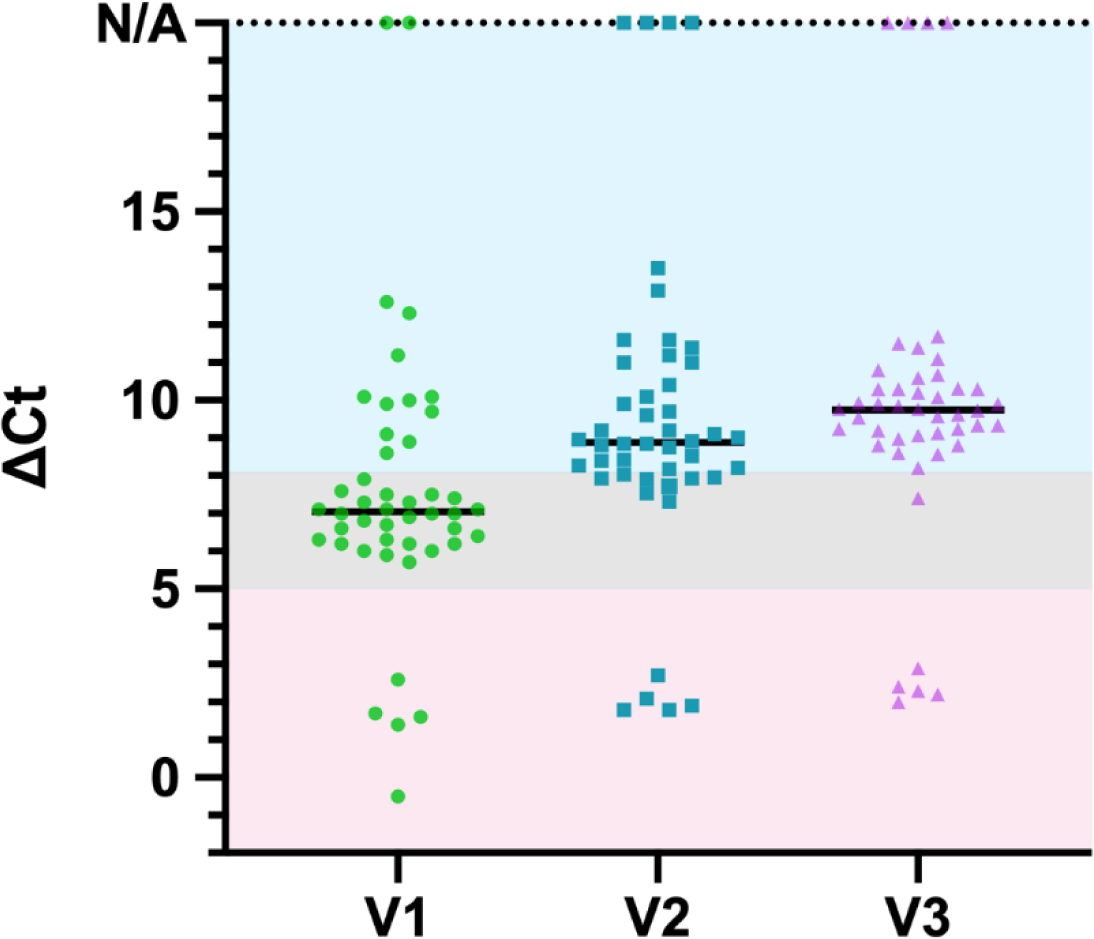
Overview of B.1.1.7 assay performance on clinical samples. Three different versions (V1, V2, V3) of B.1.1.7 primer/probe sets that varied according to the reverse primer (V1, V2, and V3 use reverse primers R14, R23, and R36, respectively) were directly compared on a selected panel of 46 SARS-CoV-2 positive clinical samples, some of which were confirmed B.1.1.7 and B.1.258Δ variants by sequencing. ΔCt values correspond to B.1.1.7 assay Ct − SARS-CoV-2 S gene assay Ct. Coloured boxes within the plot define boundaries for corresponding variant interpretation, red (ΔCt ±5) for B.1.1.7, blue (ΔCt 8-20) for ΔH69/ΔV70, grey (ΔCt 5-8) for inconclusive samples. N/A represents samples which were detected only in SARS-CoV-2 S gene assay and therefore are interpreted as consensus SARS-CoV-2.

**Supplementary Table S1.** Origin and genomic characterization of GISAID sequences used for alignment and primer/probe design.

| Country of origin | Sequences | Omitted | No deletions | ΔH69/ΔV70 only | ΔY144 only | ΔH69/ΔV70 and ΔY144 |
| --- | --- | --- | --- | --- | --- | --- |
| **Australia** | 21 | 0 | 20 (95.2 %) | 0 | 0 | 1 (4.8 %) |
| **Denmark** | 107 | 0 | 99 (92.5 %) | 8 (7.5 %) | 0 | 0 |
| **UK** | 965 | 5 | 725 (75.5 %) | 8 (0.8 %) | 0 | 227 (23.7 %) |
| **New Zealand** | 13 | 0 | 13 | 0 | 0 | 0 |
| **Sweden** | 2 | 0 | 2 | 0 | 0 | 0 |
| **Thailand** | 3 | 0 | 3 | 0 | 0 | 0 |
| **USA** | 25 | 0 | 25 | 0 | 0 | 0 |
| **Total** | 1136 | 5 | 887 | 16 | 0 | 228 |

**Supplementary Table S2.**
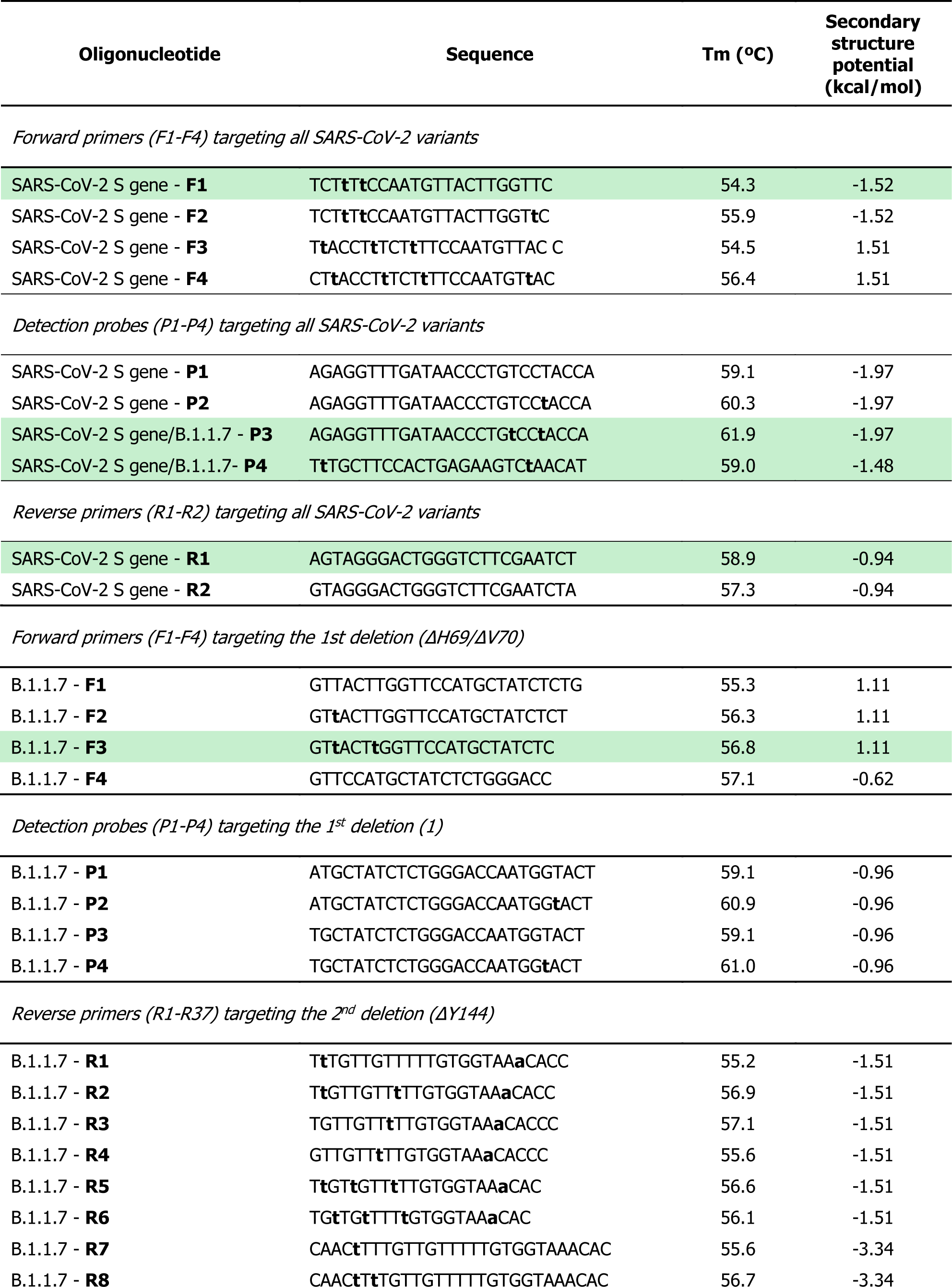
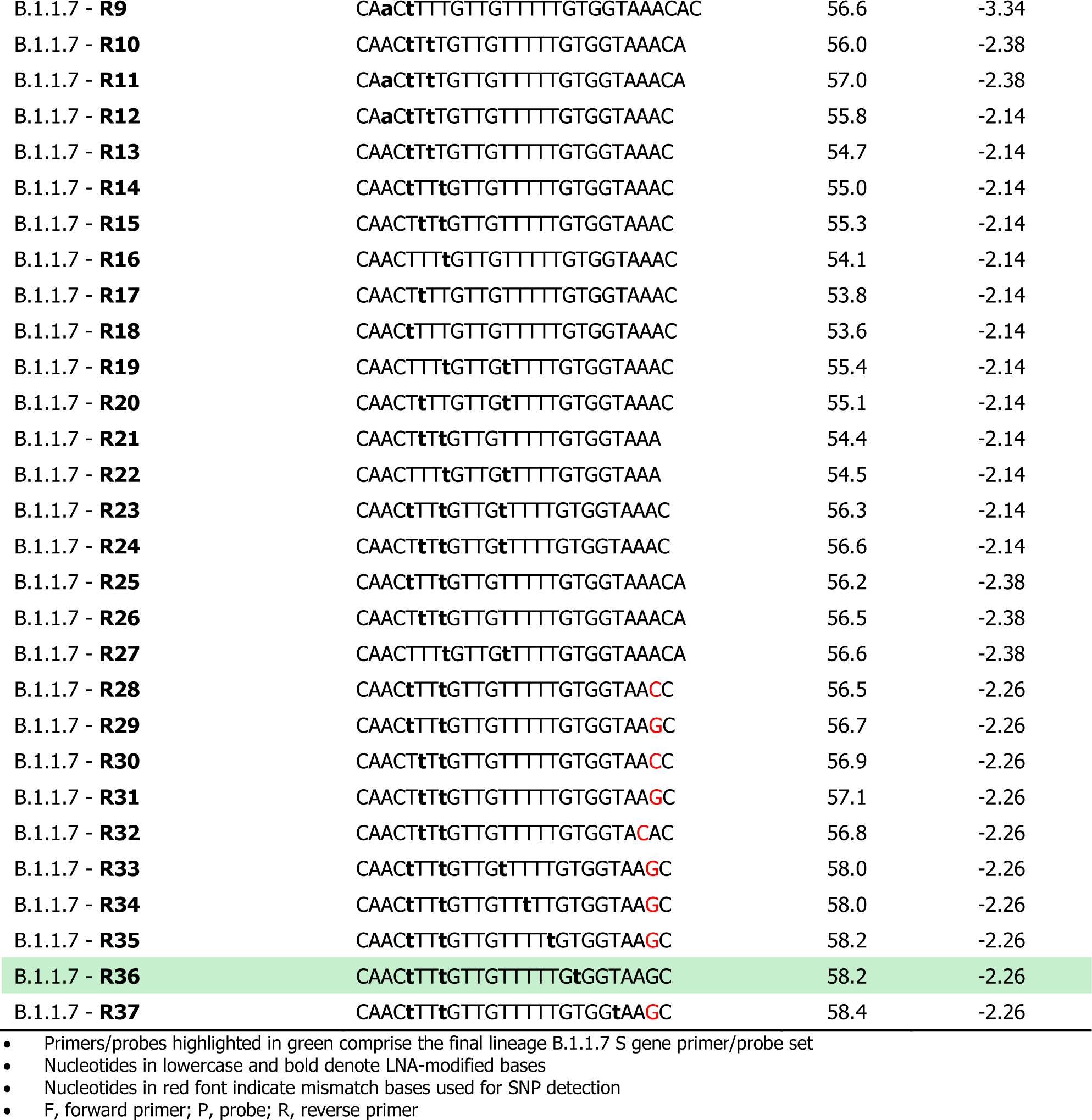
Oligonucleotide primers and probes for common SARS-CoV-2 S gene and B.1.1.7 primer/probe sets.

**Supplementary Table S3.** Assessment of clinical performance according to QUADUS-2.

| Item | Question | Answer | Domain | Risk of bias |
| --- | --- | --- | --- | --- |
| 1. | Was a consecutive or random sample of patients enrolled? | Omitted -Not applicable | Patient selection | Low |
| 2. | Was a case-control design avoided? | Yes |  |  |
| 3. | Did the study avoid inappropriate exclusions? | Yes |  |  |
| 4. | **Applicability:** Are there concerns that the included patients do not match the review question? | Low |  |  |
| 5. | Were the index test results interpreted without knowledge of the results of the reference standard? | Omitted -Not applicable | Index test | Low |
| 6. | If a threshold was used, was it prespecified? | Yes |  |  |
| 7. | **Applicability:** Are there concerns that the index test, its conduct, or its interpretation differ from the review question? | Low |  |  |
| 8. | Is the reference standard likely to correctly classify the target condition? | Yes | Reference standard | Low |
| 9. | Were the reference standard results interpreted without knowledge of the results of the index test results? | Yes |  |  |
| 10. | **Applicability:** Are there concerns that the target condition as defined by the reference standard does not match the review question? | No |  |  |
| 11. | Was there an appropriate interval between index test(s) and reference standard? | Yes | Flow and timing | Low |
| 12. | Did all patients receive a reference standard? | Yes |  |  |
| 13. | Did all patients receive the same reference standard? | Unclear |  |  |
| 14. | Were all patients included in the analysis? | Yes |  |  |

**Supplementary Table S4.**
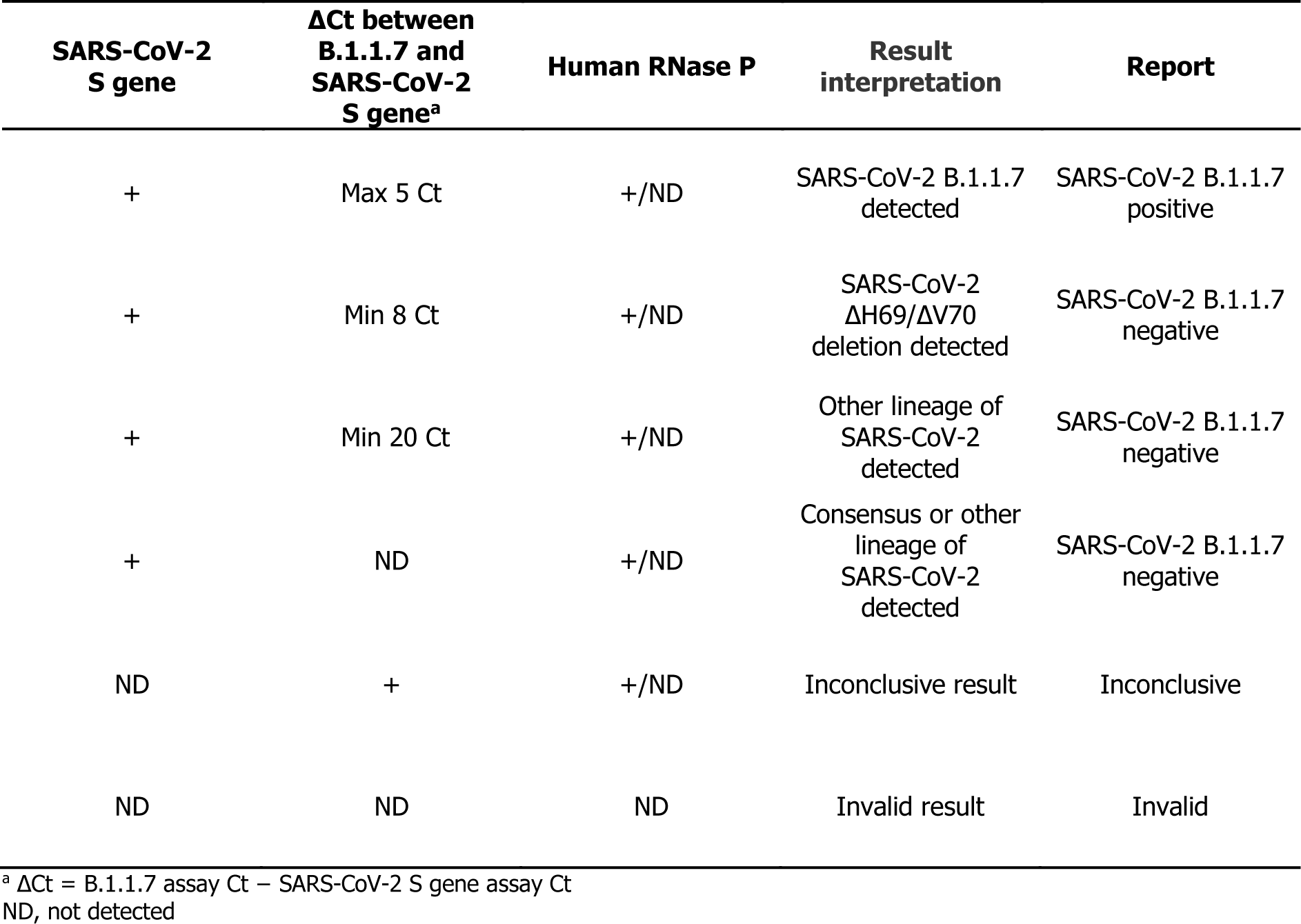
Interpretation of SARS-CoV-2 test results and corresponding actions.

**Supplementary Table S5.** Overview of clinical sample RT-qPCR results, lineage, and GISAID information.

| rTEST COVID-19 qPCR B.1.1.7 kit |  |  |  | Sequencing |  |  |
| --- | --- | --- | --- | --- | --- | --- |
| Sample ID | B.1.1.7 PCR [Ct] | S gene PCR [Ct] | ΔCt | Sequencing outcome (GISAID lineage) | Name in GISAID | Accession ID in GISAID |
| 1 | 23.8 | 23.6 | 0.2 | B.1.1.7 | UKBA-706 | EPI_ISL_875525 |
| 2 | 18.0 | 18.1 | -0.1 | B.1.1.7 | UKBA-707 | EPI_ISL_875526 |
| 3 | 33.9 | 34.5 | -0.5 | B.1.1.7 | UKBA-801 | EPI_ISL_831667 |
| 4 | 36.4 | 36.0 | 0.5 | B.1.1.7 | UKBA-802 | EPI_ISL_831668 |
| 5 | 32.3 | 31.5 | 0.8 | B.1.1.7 | UKBA-708 | EPI_ISL_875527 |
| 6 | 37.5 | 35.2 | 2.3 | B.1.1.7 | UKBA-803 | EPI_ISL_831672 |
| 7 | 23.7 | 25.0 | -1.3 | B.1.1.7 | UKBA-714 | EPI_ISL_875521 |
| 8 | 25.3 | 24.8 | 0.5 | B.1.1.7 | UKBA-713 | EPI_ISL_875520 |
| 9 | No Ct | 30.6 | - | B.1.160 | UKBA-701 | EPI_ISL_875530 |
| 10 | 28.8 | 28.5 | 0.3 | B.1.1.7 | UKBA-703 | EPI_ISL_875522 |
| 11 | 32.0 | 26.9 | 5.0 | B.1.1.7 | UKBA-705 | EPI_ISL_875524 |
| 12 | 25.2 | 22.9 | 2.3 | B.1.1.7 | UKBA-704 | EPI_ISL_875523 |
| 13 | 32.2 | 24.5 | 7.7 | B.1.258 | UKBA-702 | EPI_ISL_875528 |
| 14 | No Ct | 28.8 | - | B.1.1.243 | UKBA-715 | EPI_ISL_875516 |
| 15 | No Ct | 29.6 | - | B.1.177 | UKBA-716 | EPI_ISL_875533 |
| 16 | No Ct | 24.4 | - | B.1.177 | UKBA-717 | EPI_ISL_875534 |
| 17 | 34.3 | 29.6 | 4.7 | B.1.1.7 | UKBA-718 | EPI_ISL_875517 |
| 18 | 30.8 | 30.2 | 0.6 | B.1.1.7 | UKBA-719 | EPI_ISL_875518 |
| 19 | 25.2 | 22.6 | 2.6 | B.1.1.7 | UKBA-720 | EPI_ISL_875519 |
| 20 | 23.2 | 13.2 | 10.1 | B.1.258 | UKBA-722 | EPI_ISL_875529 |
| 21 | No Ct | 12.7 | - | B.1.1.170 | UKBA-723 | EPI_ISL_875532 |
| 22 | No Ct | 25.8 | - | B.1.1.170 | UKBA-724 | EPI_ISL_875538 |
| 23 | 21.1 | 18.8 | 2.3 | B.1.1.7 | UKBA-804 | EPI_ISL_831669 |
| 24 | 29.5 | 26.9 | 2.6 | B.1.1.7 | UKBA-805 | EPI_ISL_831670 |
| 25 | 22.8 | 20.8 | 1.9 | B.1.1.7 | UKBA-806 | EPI_ISL_831673 |
| 26 | 26.2 | 24.2 | 2.0 | B.1.1.7 | UKBA-807 | EPI_ISL_831671 |
| 27 | 21.3 | 18.6 | 2.6 | B.1.1.7 | UKBA-808 | EPI_ISL_831674 |
| 28 | 26.7 | 15.2 | 11.5 | B.1.258 | UKBA-809 | EPI_ISL_831676 |
| 29 | 30.0 | 27.8 | 2.3 | B.1.1.7 | UKBA-814 | EPI_ISL_831675 |
| 30 | 23.5 | 21.8 | 1.6 | B.1.1.7 | UKBA-815 | EPI_ISL_831663 |
| 31 | 22.9 | 18.7 | 4.1 | B.1.1.7 | UKBA-816 | EPI_ISL_831664 |
| 32 | 28.5 | 24.2 | 4.3 | B.1.1.7 | UKBA-817 | EPI_ISL_831665 |
| 33 | No Ct | 30.1 | - | B.1.258 | UKBA-818 | EPI_ISL_831666 |
| 34 | 25.4 | 24.0 | 1.4 | B.1.1.7 | UKBA-501 | EPI_ISL_779651 |
| 35 | 29.4 | 29.1 | 0.3 | B.1.1.7 | UKBA-502 | EPI_ISL_779652 |
| 36 | 30.7 | 28.9 | 1.8 | B.1.1.7 | UKBA-503 | EPI_ISL_779653 |
| 37 | 31.5 | 30.0 | 1.5 | B.1.1.7 | UKBA-504 | EPI_ISL_779654 |
| 38 | 34.3 | 29.9 | 4.4 | B.1.1.7 | UKBA-505 | EPI_ISL_779655 |
| 39 | 31.5 | 30.8 | 0.7 | B.1.1.7 | UKBA-506 | EPI_ISL_779656 |
| 40 | 37.9 | 32.2 | 5.7 | B.1.1.7 | UKBA-507 | EPI_ISL_779657 |
| 41 | 30.2 | 29.1 | 1.1 | B.1.1.7 | UKBA-508 | EPI_ISL_779658 |
| 42 | 37.6 | 34.4 | 3.2 | B.1.1.7 | UKBA-509 | EPI_ISL_779659 |
| 43 | 39.7 | 35.2 | 4.5 | B.1.1.7 | UKBA-512 | EPI_ISL_779660 |
| 44 | 25.8 | 14.7 | 11.1 | B.1.258 | UKBA-1001 | EPI_ISL_903980 |
| 45 | No Ct | 23.1 | - | B.1.1.170 | UKBA-1002 | EPI_ISL_903981 |
| 46 | 24.4 | 15.2 | 9.2 | B.1.258 | UKBA-1004 | EPI_ISL_903983 |
| 47* | 35.8 | 27.1 | 8.7 | B.1.258 | UKBA-1005 | EPI_ISL_903984 |
| 48* | 29.1 | 18.3 | 10.8 | B.1.258 | UKBA-1006 | EPI_ISL_903985 |
| 49 | 15.6 | 16.8 | -1.2 | B.1.1.7 | UKBA-1007 | EPI_ISL_903986 |
| 50 | 25.4 | 16.1 | 9.3 | B.1.258 | UKBA-1008 | EPI_ISL_903987 |
| 51 | No Ct | 16.5 | - | B.1.160 | UKBA-1009 | EPI_ISL_903988 |
| 52 | 19.9 | 17.4 | 2.5 | B.1.1.7 | UKBA-1010 | EPI_ISL_903989 |
| 53 | 27.6 | 25.5 | 2.1 | B.1.1.7 | UKBA-1011 | EPI_ISL_903990 |
| 54* | 37.1 | 26.0 | 11.1 | B.1.258 | UKBA-1012 | EPI_ISL_903991 |
| 55 | No Ct | 15.1 | - | B.1.177 | UKBA-1013 | EPI_ISL_903992 |
| 56 | 28.0 | 19.6 | 8.4 | B.1.258 | UKBA-1014 | EPI_ISL_903993 |
| 57 | No Ct | 23.4 | - | B.1.1.277 | UKBA-1015 | EPI_ISL_903994 |
| 58* | 35.5 | 26.9 | 8.6 | B.1.258 | UKBA-1016 | EPI_ISL_903995 |
| 59 | No Ct | 16.3 | - | B.1.160 | UKBA-1017 | EPI_ISL_903996 |
| 60 | 33.2 | 22.9 | 10.3 | B.1.258 | UKBA-1018 | EPI_ISL_903997 |
| 61 | 26.5 | 17.5 | 9.0 | B.1.258 | UKBA-1020 | EPI_ISL_903999 |
| 62 | 19.8 | 20.7 | -0.9 | B.1.1.7 | UKBA-1021 | EPI_ISL_904000 |
| 63 | No Ct | 18.9 | - | B.1.221 | UKBA-1022 | EPI_ISL_904001 |
| 64* | 35.2 | 25.3 | 9.9 | B.1.258 | UKBA-1023 | EPI_ISL_904002 |
| 65* | No Ct | 28.9 | - | B.1.258 | UKBA-1024 | EPI_ISL_904003 |
| 66 | 21.7 | 19.4 | 2.4 | B.1.1.7 | UKBA-1101 | EPI_ISL_959643 |
| 67 | No Ct | 17.1 | - | B.1.160 | UKBA-1102 | EPI_ISL_959642 |
| 68 | 25.8 | 16.4 | 9.4 | B.1.258 | UKBA-1103 | EPI_ISL_959648 |
| 69 | 17.7 | 15.3 | 2.4 | B.1.1.7 | UKBA-1104 | EPI_ISL_959645 |
| 70 | No Ct | 17.9 | - | B.1.1.170 | UKBA-1105 | EPI_ISL_959647 |
| 71 | 21.7 | 18.9 | 2.9 | B.1.1.7 | UKBA-1106 | EPI_ISL_959646 |
| 72 | 26.5 | 22.6 | 3.9 | B.1.1.7 | UKBA-1107 | EPI_ISL_959644 |
| 73 | 32.3 | 20.6 | 11.6 | B.1.258 | UKBA-1108 | EPI_ISL_959649 |
| 74 | No Ct | 25.0 | - | B.1.1.7 | UKBA-1109 | EPI_ISL_959637 |
| 75* | 33.8 | 30.3 | 3.5 | B.1.1.7 | UKBA-1110 | EPI_ISL_959638 |
| 76* | 21.0 | 19.5 | 1.5 | B.1.1.7 | UKBA-1111 | EPI_ISL_959639 |
| 77* | 26.3 | 23.8 | 2.5 | B.1.1.7 | UKBA-1112 | EPI_ISL_959640 |
| 78 | No Ct | 23.2 | - | B.1.1.170 | UKBA-1113 | EPI_ISL_959641 |
| 79 | 29.9 | 27.5 | 2.5 | B.1.1.7 | UKBA-1114 | EPI_ISL_959627 |
| 80 | 32.7 | 24.2 | 8.5 | B.1.258 | UKBA-1115 | EPI_ISL_959630 |
| 81 | 31.0 | 28.8 | 2.1 | B.1.1.7 | UKBA-1116 | EPI_ISL_959628 |
| 82 | 25.3 | 23.0 | 2.3 | B.1.1.7 | UKBA-1117 | EPI_ISL_959626 |
| 83 | 26.9 | 16.7 | 10.2 | B.1.258 | UKBA-1118 | EPI_ISL_959631 |
| 84 | 20.9 | 17.5 | 3.4 | B.1.1.7 | UKBA-1119 | EPI_ISL_959629 |
| 85 | 23.8 | 19.0 | 4.8 | B.1.1.7 | UKBA-1120 | EPI_ISL_959632 |
| 86 | 21.6 | 19.3 | 2.2 | B.1.1.7 | UKBA-1121 | EPI_ISL_959633 |
| 87 | 19.0 | 16.3 | 2.7 | B.1.1.7 | UKBA-1122 | EPI_ISL_959634 |
| 88 | 19.4 | 17.2 | 2.1 | B.1.1.7 | UKBA-1123 | EPI_ISL_959635 |
| 89 | 28.0 | 16.7 | 11.4 | B.1.258 | UKBA-1124 | EPI_ISL_959636 |
| 90 | 18.1 | 16.7 | 1.4 | B.1.1.7 | UKBA-1207 | EPI_ISL_959604 |
| 91 | 26.4 | 22.8 | 3.6 | B.1.1.7 | UKBA-1208 | EPI_ISL_959605 |
| 92 | 15.5 | 13.3 | 2.3 | B.1.1.7 | UKBA-1209 | EPI_ISL_959606 |
| 93 | 23.8 | 14.1 | 9.7 | B.1.258 | UKBA-1210 | EPI_ISL_959607 |
| 94 | 18.4 | 16.9 | 1.5 | B.1.1.7 | UKBA-1211 | EPI_ISL_959608 |
| 95 | 17.2 | 15.3 | 1.8 | B.1.1.7 | UKBA-1212 | EPI_ISL_959609 |
| 96 | 22.8 | 21.5 | 1.3 | B.1.1.7 | UKBA-1213 | EPI_ISL_959610 |
| 97 | 16.4 | 14.4 | 2.0 | B.1.1.7 | UKBA-1214 | EPI_ISL_959611 |
| 98 | 18.9 | 16.9 | 2.1 | B.1.1.7 | UKBA-1215 | EPI_ISL_959612 |
| 99 | 14.9 | 13.4 | 1.5 | B.1.1.7 | UKBA-1216 | EPI_ISL_959613 |
| 100 | 16.1 | 14.4 | 1.8 | B.1.1.7 | UKBA-1217 | EPI_ISL_959614 |
| 101 | 24.3 | 22.0 | 2.3 | B.1.1.7 | UKBA-1218 | EPI_ISL_959615 |
| 102 | 17.6 | 15.3 | 2.3 | B.1.1.7 | UKBA-1219 | EPI_ISL_959616 |
| 103 | 20.2 | 17.8 | 2.4 | B.1.1.7 | UKBA-1221 | EPI_ISL_959617 |
| 104 | 15.9 | 14.1 | 1.8 | B.1.1.7 | UKBA-1222 | EPI_ISL_959618 |
| 105 | No Ct | 23.9 | - | B.1.258 | UKBA-1223 | EPI_ISL_959619 |
| 106 | 27.1 | 16.9 | 10.2 | B.1.258 | UKBA-1224 | EPI_ISL_959620 |

\* Samples with an asterisk were re-tested due to an inconclusive result in the first test. The results depict the re-test values.

**Supplementary Table S6.** Surveillance of lineage B.1.1.7 prevalence in the Slovak Republic.

| Region | B.1.1.7<br>($\Delta H69/\Delta V70 + \Delta Y144$) | $\Delta H69/\Delta V70$<br>only | Other<br>lineage<br>of<br>SARS-CoV-2 | Inconclusive | Sample<br>size | B.1.1.7<br>prevalence |
| --- | --- | --- | --- | --- | --- | --- |
| *Retest date: 2 February 2021* |  |  |  |  |  |  |
| **Banská Bystrica** | 132 | 1 | 46 | 4 | 183 | 74% |
| **Bratislava** | 306 | 12 | 73 | 4 | 395 | 78% |
| **Košice** | 163 | 0 | 65 | 12 | 240 | 71% |
| **Nitra** | 144 | 5 | 51 | 5 | 205 | 72% |
| **Prešov** | 124 | 3 | 66 | 3 | 196 | 64% |
| **Trenčín** | 131 | 5 | 25 | 6 | 167 | 81% |
| **Trnava** | 357 | 6 | 57 | 17 | 437 | 85% |
| **Žilina** | 69 | 9 | 54 | 7 | 139 | 52% |
| **Total** | **1426** | **41** | **437** | **58** | **1962** | **75%** |
| *Retest date: 17 February 2021* |  |  |  |  |  |  |
| **Banská Bystrica** | 242 | 0 | 156 | 4 | 402 | 61% |
| **Bratislava** | 217 | 0 | 46 | 4 | 267 | 83% |
| **Košice** | 235 | 1 | 114 | 18 | 368 | 67% |
| **Nitra** | 101 | 0 | 45 | 5 | 151 | 69% |
| **Prešov** | 145 | 3 | 45 | 27 | 220 | 75% |
| **Trenčín** | 248 | 0 | 50 | 0 | 298 | 83% |
| **Trnava** | 348 | 0 | 53 | 17 | 418 | 87% |
| **Žilina** | 174 | 0 | 84 | 0 | 258 | 67% |
| **Total** | **1710** | **4** | **593** | **75** | **2382** | **74%** |
| *Retest date: 3 March 2021* |  |  |  |  |  |  |
| **Banská Bystrica** | 211 | 0 | 39 | 16 | 266 | 84% |
| **Bratislava** | 222 | 1 | 22 | 11 | 256 | 91% |
| **Košice** | 225 | 0 | 82 | 60 | 367 | 73% |
| **Nitra** | 128 | 2 | 21 | 16 | 167 | 85% |
| **Prešov** | 206 | 4 | 54 | 44 | 308 | 78% |
| **Trenčín** | 322 | 7 | 51 | 32 | 412 | 85% |
| **Trnava** | 349 | 6 | 35 | 19 | 409 | 89% |
| **Žilina** | 262 | 1 | 90 | 4 | 357 | 74% |
| **Total** | **1925** | **21** | **394** | **202** | **2542** | **82%** |

